# Depressive symptoms and viral load suppression among adolescents and youth living with HIV in Malawi: a cross-sectional analysis of MPHIA 2020–2021

**DOI:** 10.64898/2026.10.05.26364733

**Authors:** Grace Ndhlovu, Alex de Voux

**Affiliations:** Division of Epidemiology and Biostatistics, School of Public Health, University of Cape Town, Cape Town, South Africa

**Keywords:** adolescents and youth living with HIV, viral load suppression, depressive symptoms, PHQ-2, HIV-status awareness, antiretroviral therapy, Malawi, population-based survey

## Abstract

**Background:** Viral load suppression (VLS) remains suboptimal among adolescents and youth living with HIV (AYLWH) in Malawi. Depressive symptoms may affect treatment adherence and HIV outcomes, but evidence is limited. We assessed the association between depressive symptoms and VLS among AYLWH using Malawi Population-based HIV Impact Assessment (MPHIA) 2020–2021 data.

**Methods:** We conducted a cross-sectional secondary analysis of AYLWH aged 15–24 years. VLS was defined as HIV RNA less than 200 copies/mL and depressive symptoms as a Patient Health Questionnaire-2 score of 3 or higher. Survey-weighted logistic regression assessed associations with VLS, adjusting for sex, education, residence, household wealth, alcohol use, Teen Club participation, and awareness of HIV-positive status.

**Findings:** Among 201 participants, viral-load results were available for 200; 137 (68.5%) achieved VLS. Six (3.0%) participants had depressive symptoms. Depressive symptoms were not associated with VLS in univariable analysis (OR 0.8, 95% CI 0.1–4.6; p=0.80) or multivariable analysis (aOR 0.3, 95% CI 0.1–2.4; p=0.28). Urban residence (aOR 0.2, 95% CI 0.1–0.5; p=0.0020) and unawareness of HIV-positive status (aOR 0.1, 95% CI 0.02–0.1; p<0.001) were associated with lower odds of VLS.

**Interpretation:** Depressive symptoms were not significantly associated with VLS, although their low prevalence and PHQ-2 limitations reduced precision. The findings highlight the importance of strengthening HIV-status awareness and addressing context-specific barriers to care among AYLWH.

**Funding:** This secondary analysis received no specific funding.

## Introduction

HIV viral load suppression (VLS) is a cornerstone of HIV management because it improves individual health outcomes, reduces HIV transmission, and supports progress towards the UNAIDS 95-95-95 targets ^1-3^. In Malawi, national VLS, defined in Malawi Population-based HIV Impact Assessment (MPHIA) 2020–2021 as HIV RNA <1000 copies/mL, increased from 68% in 2015–2016 to 87% in 2020–2021 ^1,2^. Among those receiving antiretroviral therapy (ART), VLS has reached 97%, meeting the UNAIDS target ^1,2^. However, these national gains mask persistent gaps among adolescents and youth living with HIV (AYLWH), particularly those aged 15–24 years ^1,4,5^. MPHIA 2020–2021 reports adolescents and youth accounted for a substantial proportion of new HIV infections, yet only 70% had detectable antiretroviral drugs and 74% achieved VLS ^1,2^. These figures fall far short of the UNAIDS 95% target for key population groups, highlighting an urgent need for targeted interventions to address the unique challenges faced by this vulnerable group and bridge the gap in epidemic control efforts.

Suboptimal VLS among AYLWH is influenced by biological, behavioural, psychosocial, and structural factors that affect ART adherence and retention in care ^4-8^. Among these, mental health, particularly depression, is increasingly recognised as an important but often underaddressed determinant of HIV outcomes. Depressive symptoms may reduce motivation, self-efficacy, and treatment engagement, thereby undermining consistent ART use ^5,6,8-11^. This study focuses on depressive symptoms rather than a clinical diagnosis of depression. Depressive symptoms may also contribute indirectly to poor HIV treatment outcomes through behaviours such as substance use and risky sexual activities ^6,8,12^. In addition, evidence suggests that depression-related biological mechanisms, including inflammatory responses, may contribute to poorer immune outcomes among people living with HIV^9,13^.

Evidence on the burden and effects of depressive symptoms among AYLWH in sub-Saharan Africa remains variable. A systematic review reported that depressive symptoms among HIV-positive adolescents in sub-Saharan Africa range from 4% to 53%, while major depressive disorder has been estimated at 16% to 40%, partly reflecting differences in study populations and screening tools^14^. Studies examining the association between depression and VLS have also produced mixed findings. A study among youth in rural South Africa, which assessed depressive symptoms using the 20-item Center for Epidemiological Studies Depression (CES-D) scale, found no significant association with viral non-suppression, whereas a South African private-sector cohort, in which depression was identified from recorded ICD-10 diagnoses, reported a higher risk of viral non-suppression among individuals with depression ^6,10^. In Malawi, available studies have reported depression prevalence ranging from 19% to 22%, depending on the population studied and the assessment tool used ^15,16^. Evidence from Malawi also suggests that depressive symptoms are associated with poor ART adherence among AYLWH, which could in turn affect VLS ^5^. However, recent national-level evidence on depressive symptoms and their association with VLS among AYLWH in Malawi remains limited.

The integration of mental health services into HIV care in Malawi is still developing. National ART programmes and adolescent-focused interventions, such as Teen Clubs, structured peer-support programmes for adolescents and youth living with HIV, have improved aspects of HIV care among adolescents ^5,17^. However, the integration of mental health services into HIV care remains inadequate ^18^. In 2022, Malawi introduced mental health screening into the national HIV clinical management guidelines, particularly for priority clients such as those newly diagnosed with HIV, those with high viral loads (≥1,000 copies/mL), and those returning to care after treatment interruptions^19^. However, implementation remains limited, and many adolescents and youth who may benefit from mental health assessment and support may not be routinely reached^18^. Persistent stigma, limited mental health infrastructure, and shortages of trained providers further constrain the identification and management of depressive symptoms within HIV care services^5,17,20,21^.

Beyond mental health, socioeconomic and structural factors may also shape VLS among AYLWH. Residence, education, wealth, alcohol use, psychosocial support, and awareness of HIV status may influence ART access, adherence, and continuity of care^4-6,22^. In Malawi, rural and urban settings present different barriers to HIV care. Rural communities may face challenges related to poverty, transport, and distance to services, while urban areas may be affected by overcrowding, informal settlements, economic insecurity, and limited youth-friendly service access^23-25^. Factors such as sex, education level, substance use, and psychosocial support further interact with these contextual inequalities, shaping HIV outcomes in varying ways ^4,12,26^. Awareness of HIV status is also critical, as it enables timely linkage to care and ART initiation ^1,3^. However, a significant proportion of AYLWH remain unaware of their HIV status, increasing the risk of delayed treatment, viral non-suppression, and onward transmission^1,6^.

These interrelated influences were considered within Social Action Theory (SAT), which suggests that health outcomes are shaped by the interaction between contextual factors, self-regulation, and health-related behaviours^4,5,7^. Among AYLWH, depressive symptoms, socioeconomic conditions, HIV status awareness, and access to psychosocial support may influence motivation, coping, ART adherence, and engagement with care, ultimately affecting VLS. This framework therefore guided the selection of individual, behavioural, and structural factors examined in relation to VLS.

This study uses MPHIA 2020–2021 data to examine the association between depressive symptoms and VLS among AYLWH. Guided by SAT, the analysis examines associations between depressive symptoms, behavioural and structural factors, and viral suppression. The findings may inform targeted mental health and HIV care interventions to improve treatment outcomes and support Malawi’s efforts towards ending the HIV epidemic.

## Methods

### Study design and data source

We conducted a cross-sectional secondary analysis of data from the Malawi Population-based HIV Impact Assessment (MPHIA) 2020–2021. MPHIA 2020–2021 was a nationally representative household survey designed to assess HIV-related indicators among individuals aged 15 years and older in Malawi ^1^. The present analysis focused on adolescents and youth living with HIV (AYLWH) aged 15–24 years.

### Study population and sampling

MPHIA 2020–2021 used a two-stage stratified cluster sampling design. In the first stage, 438 enumeration areas were selected from the Malawi 2018 census sampling frame using probability proportional to size and stratification by seven health zones^1^. In the second stage, households were randomly selected within each enumeration area, with an average of 35 households selected per cluster^1^.

Individuals aged 15 years and older who had stayed in a selected household the previous night were eligible to participate. Of 13,958 eligible households, 12,815 (91.8%) completed the household interview. Overall, 30,049 individuals were eligible to participate, 26,519 (88.3%) completed individual interviews, and 22,662 (85.5%) consented to blood collection^1^. Among participants with HIV test results, 2,464 were living with HIV, of whom 201 were aged 15–24 years and were included in this secondary analysis. Viral-load results were available for 200 participants. No separate sample-size calculation was undertaken for this secondary analysis; all eligible participants aged 15–24 years living with HIV in the MPHIA 2020– 2021 dataset were included.

### Data collection and laboratory procedures

MPHIA 2020–2021 data collection was conducted from January, 2020, to April, 2021, with a temporary pause from April, 2020, to March, 2021, because of the COVID-19 pandemic^1^. Trained field teams administered standardised electronic questionnaires using mobile tablet devices. Questionnaires were available in English, Chichewa, and Tumbuka and collected sociodemographic, behavioural, health, and psychosocial information ^1^.

Blood samples were collected from consenting participants and processed for HIV testing, viral-load measurement, and antiretroviral detection according to MPHIA survey procedures^1^. HIV viral load was measured using the COBAS AmpliPrep/TaqMan assay, with dried blood spot specimens used when plasma specimens were unavailable^1^.

### Study variables

The primary outcome was viral load suppression (VLS), defined as HIV RNA less than 200 copies/mL and categorised as suppressed or not suppressed ^3^. Participants without a viral-load result were excluded from analyses in which VLS was the outcome.

The primary exposure was depressive symptoms, assessed using the Patient Health Questionnaire-2 (PHQ-2). The PHQ-2 assesses loss of interest or pleasure and feelings of being down, depressed, or hopeless during the preceding 2 weeks. Each item is scored from 0 to 3, giving a total score ranging from 0 to 6. A PHQ-2 score of 3 or higher was used to indicate the presence of depressive symptoms ^27^. The PHQ-2 is an abbreviated two-item version of the PHQ-9 and is intended as a brief screening instrument rather than a diagnostic assessment. At this cutoff, it demonstrated a sensitivity of 0.72 and specificity of 0.85 among adults living with HIV in western Kenya ^27^. The PHQ-2 has not been formally validated among Malawian adolescents and youth, and its brevity may limit its sensitivity compared with more comprehensive instruments.

Covariates were selected *a priori* on the basis of SAT and published evidence on factors associated with VLS among AYLWH. Sex was recorded as male or female. Age was recorded in completed years and restricted to 15–24 years for this analysis. Place of residence was classified as urban or rural according to the MPHIA enumeration-area classification. Education represented the highest level of formal education completed and was categorised as no education, primary, secondary, or more than secondary education for descriptive analyses. Household wealth was represented by the MPHIA-derived wealth quintile, derived from household assets, housing materials, and access to utilities, and categorised from lowest to highest. Health zone represented the seven MPHIA sampling zones: North, Central East, Central West, South East, South West, Lilongwe City, and Blantyre City. Alcohol use reflected self-reported alcohol consumption. Awareness of HIV-positive status indicated whether participants knew their HIV-positive status before receiving the survey test result. Participation in psychosocial interventions referred to participation in Teen Club, a structured peer-support programme for adolescents and youth living with HIV ^17^.

### Statistical analysis

De-identified MPHIA 2020–2021 data were analysed using R version 4.3. Participant characteristics were summarised using frequencies and percentages for categorical variables and medians with interquartile ranges for continuous variables.

Survey-weighted analyses accounted for the complex two-stage stratified cluster sampling design of MPHIA 2020–2021. A survey design object was created using the survey package in R ^28^, incorporating primary sampling units, sampling strata, and trimmed blood-test non-response-adjusted weights from the MPHIA intermediary person weights file. Survey weights accounted for unequal probabilities of selection and differential non-response, and strata and clusters were incorporated to obtain appropriate variance estimates and standard errors.

Weighted logistic regression was used to assess the association between depressive symptoms and VLS among the 200 participants with available viral-load results. Univariable logistic regression was used to estimate crude associations, followed by multivariable weighted logistic regression to estimate adjusted associations. Covariates for the multivariable model were selected a priori on the basis of the conceptual framework and published evidence on factors associated with VLS among AYLWH ^4-6,8^. The multivariable model included depressive symptoms, sex, place of residence, education level, household wealth quintile, alcohol use, awareness of HIV-positive status, and participation in psychosocial interventions. For regression analyses, education was dichotomised as basic education (no education or primary education) versus higher education (secondary or more than secondary education). Alcohol use was dichotomised as any reported alcohol use versus no alcohol use, with participants who responded “don’t know/refused” included in the no-use category. Teen Club participation was dichotomised as yes versus no, with participants who responded “don’t know/refused” included in the no category. Associations were reported as odds ratios (ORs) and adjusted odds ratios (aORs) with 95% CIs. Statistical significance was assessed at p<0.05.

### Ethics

The primary MPHIA 2020–2021 survey received ethical approval from the National Health Sciences Research Committee of Malawi, the United States Centers for Disease Control and Prevention, and the Columbia University institutional review boards ^1^. Participants aged 18 years and older provided informed consent. For participants aged 15–17 years, parental or guardian permission and participant assent were obtained. Consent procedures were conducted in participants’ preferred language ^1^.

This secondary analysis used existing de-identified data, and no new participant data were collected. Ethical approval for the secondary analysis was obtained from the University of Cape Town Faculty of Health Sciences Human Research Ethics Committee (HREC ref 671/2024).

## Results

### Participant characteristics

The final analytical sample comprised 201 adolescents and youth living with HIV aged 15–24 years. Viral load results were available for 200 participants; one participant had no viral-load result. Among participants with available viral-load results, 137 of 200 (68.5%) achieved viral load suppression (VLS) and 63 of 200 (31.5%) did not.

The median age was 21 years (IQR 18–23). Overall, 150 of 201 (74.6%) participants were female, 128 (63.7%) lived in rural areas, and 142 (70.7%) had attained primary-level education. Most participants (153 [76.1%]) were aware of their HIV-positive status before receiving the survey test result. Ten (4.9%) participants reported participation in Teen Club, and six (3.0%) had depressive symptoms.

Among participants with VLS, 96 of 137 (70.1%) lived in rural areas and 124 (90.5%) were aware of their HIV-positive status. Among participants without VLS, 32 of 63 (50.8%) lived in urban areas and 35 (55.6%) were unaware of their HIV-positive status. Depressive symptoms were uncommon in both groups, occurring in four of 137 (2.9%) participants with VLS and two of 63 (3.2%) participants without VLS (Table 1).

**Table 1:** Unweighted demographic and clinical characteristics of study participants, overall and by viral load suppression status.

| Characteristic | Overall (n=201) | Viral load suppression (n=137) | Viral load non-suppression (n=63) |
| --- | --- | --- | --- |
| Age (years), median (IQR) | 21 (18–23) | 21 (18–23) | 20 (18.5–22) |
| **Sex, n (%)** |  |  |  |
| Male | 51 (25.4) | 35 (25.6) | 16 (25.4) |
| Female | 150 (74.6) | 102 (74.5) | 47 (74.6) |
| **Type of residence, n (%)** |  |  |  |
| Rural | 128 (63.7) | 96 (70.1) | 31 (49.2) |
| Urban | 73 (36.3) | 41 (29.9) | 32 (50.8) |
| **Highest level of education, n (%)** |  |  |  |
| No education | 2 (1.0) | 0 | 2 (3.2) |
| Primary | 142 (70.7) | 102 (74.5) | 40 (63.5) |
| Secondary | 54 (26.9) | 34 (24.8) | 19 (30.2) |
| More than secondary | 3 (1.5) | 1 (0.7) | 2 (3.2) |
| **Wealth quintile, n (%)** |  |  |  |
| Lowest | 30 (14.9) | 22 (16.1) | 8 (12.7) |
| Second | 22 (10.9) | 17 (12.4) | 5 (7.9) |
| Middle | 43 (21.4) | 30 (21.9) | 13 (20.6) |
| Fourth | 49 (24.4) | 36 (26.3) | 12 (19.1) |
| Highest | 57 (28.4) | 32 (23.4) | 25 (39.7) |
| **Health zone, n (%)** |  |  |  |
| Blantyre City | 34 (16.9) | 20 (14.6) | 14 (22.2) |
| South East | 50 (24.9) | 32 (23.4) | 18 (28.6) |
| South West | 68 (33.8) | 51 (37.2) | 17 (26.9) |
| Lilongwe City | 19 (9.5) | 8 (5.8) | 11 (17.5) |
| Central East | 6 (2.9) | 4 (2.9) | 1 (1.6) |
| Central West | 11 (5.5) | 11 (8.0) | 0 |
| North | 13 (6.5) | 11 (8.0) | 2 (3.2) |
| **Alcohol use, n (%)** |  |  |  |
| Yes | 22 (10.9) | 12 (8.8) | 10 (15.9) |
| No | 178 (88.7) | 124 (90.5) | 53 (84.1) |
| Don't know/refused | 1 (0.5) | 1 (0.7) | 0 |
| **HIV-status awareness, n (%)** |  |  |  |
| Aware | 153 (76.1) | 124 (90.5) | 28 (44.4) |
| Unaware | 48 (23.9) | 13 (9.5) | 35 (55.6) |
| **Teen Club participation, n (%)** |  |  |  |
| Yes | 10 (4.9) | 7 (5.1) | 3 (4.8) |
| No | 182 (90.6) | 125 (91.2) | 56 (88.9) |
| Don't know/refused | 9 (4.5) | 5 (3.7) | 4 (6.4) |
| **Depressive symptoms, n (%)** |  |  |  |
| Present | 6 (3.0) | 4 (2.9) | 2 (3.2) |
| Absent | 195 (97.0) | 133 (97.1) | 61 (96.8) |

IQR=interquartile range; VLS=viral load suppression. One of the 201 participants had no viral-load result and is included in the overall column but not in the VLS-stratified columns. Percentages in the VLS and viral load non-suppression columns use n=137 and n=63 as denominators, respectively.

### Survey-weighted participant characteristics

After weighting, participant characteristics remained broadly similar to the unweighted analysis (Table 2). Urban residence increased slightly from 36.3% to 37.1%, primary education decreased from 70.7% to 69.5%, secondary education increased from 26.9% to 27.7%, and reported alcohol use increased from 10.9% to 11.2%. Among participants with VLS, awareness of HIV-positive status decreased marginally from 90.5% to 90.0%.

**Table 2:** Survey-weighted demographic and clinical characteristics of study participants, overall and by viral load suppression status.

| Characteristic | Overall weighted estimate | VLS weighted estimate | Viral load non-suppression weighted estimate |
| --- | --- | --- | --- |
| Age (years), mean | 20.3 | 20.3 | 20.2 |
| **Sex, %** |  |  |  |
| Male | 29.2 | 29.7 | 28.4 |
| Female | 70.8 | 70.3 | 71.6 |
| **Type of residence, %** |  |  |  |
| Rural | 62.9 | 70.1 | 45.9 |
| Urban | 37.1 | 29.9 | 54.1 |
| **Highest level of education, %** |  |  |  |
| No education | 1.0 | 0 | 3.2 |
| Primary | 69.5 | 73.4 | 62.0 |
| Secondary | 27.7 | 25.8 | 30.7 |
| More than secondary | 1.8 | 0.8 | 4.1 |
| **Wealth quintile, %** |  |  |  |
| Lowest | 14.6 | 15.9 | 11.8 |
| Second | 11.0 | 12.9 | 6.9 |
| Middle | 21.2 | 21.9 | 20.0 |
| Fourth | 24.8 | 26.4 | 19.9 |
| Highest | 28.5 | 23.1 | 41.3 |
| **Health zone, %** |  |  |  |
| Blantyre City | 16.5 | 13.7 | 23.4 |
| South East | 22.6 | 20.8 | 27.0 |
| South West | 30.5 | 33.4 | 24.2 |
| Lilongwe City | 10.3 | 6.1 | 20.0 |
| Central East | 3.2 | 3.1 | 1.8 |
| Central West | 8.7 | 12.6 | 0 |
| North | 8.2 | 10.3 | 3.6 |
| **Alcohol use, %** |  |  |  |
| Yes | 11.2 | 8.7 | 17.1 |
| No | 88.2 | 90.4 | 82.9 |
| Don't know/refused | 0.6 | 0.9 | 0 |
| **HIV-status awareness, %** |  |  |  |
| Aware | 76.1 | 90.0 | 44.1 |
| Unaware | 23.9 | 10.0 | 55.9 |
| **Teen Club participation, %** |  |  |  |
| Yes | 5.1 | 5.4 | 4.4 |
| No | 90.7 | 91.4 | 89.0 |
| Don't know/refused | 4.2 | 3.2 | 6.6 |
| **Depressive symptoms, %** |  |  |  |
| Present | 2.6 | 2.5 | 3.1 |
| Absent | 97.4 | 97.5 | 97.0 |

Values are survey-weighted means for age and survey-weighted percentages for categorical variables. One participant without a viral-load result was excluded from VLS-stratified estimates.

### Factors associated with viral load suppression

Survey-weighted logistic regression analyses included the 200 participants with available viral-load results (Table 3). Depressive symptoms were not associated with VLS in either the univariable analysis (OR 0.8, 95% CI 0.1–4.6; p=0.80) or the multivariable analysis (aOR 0.3, 95% CI 0.1–2.4; p=0.28).

**Table 3:**
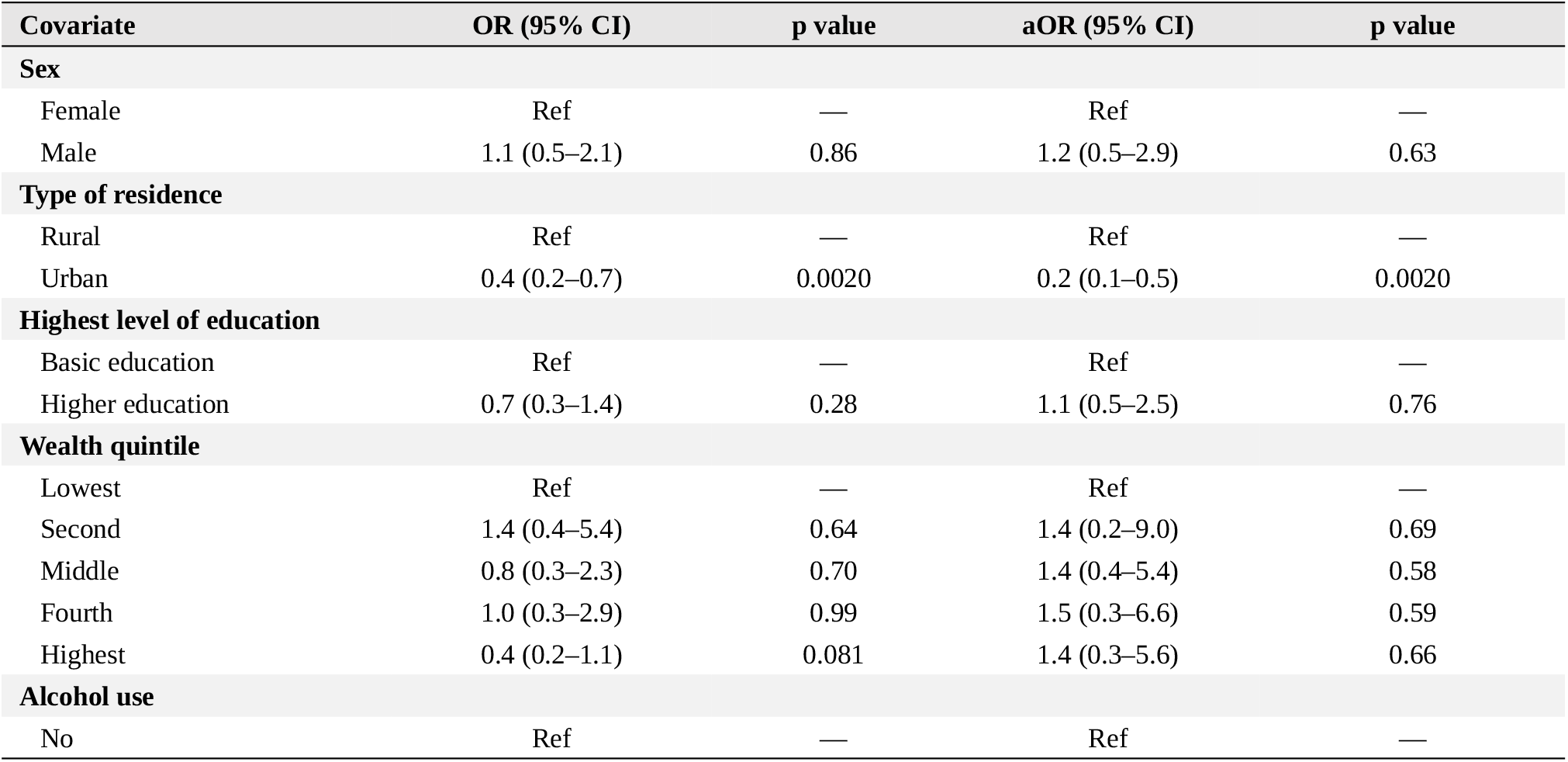
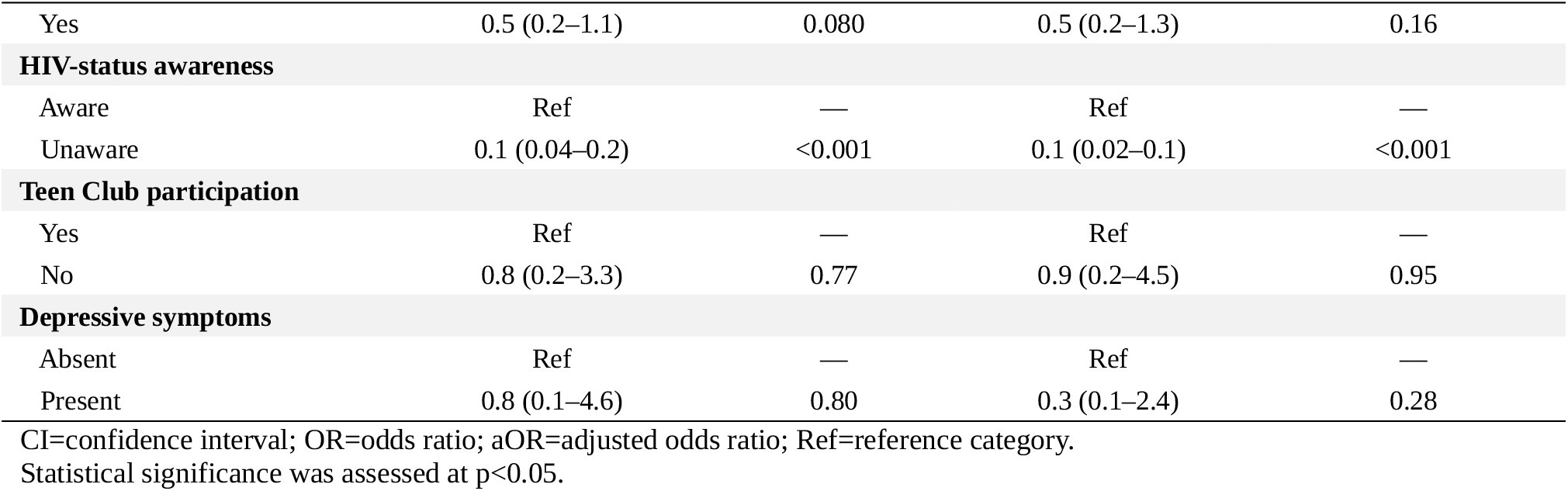
Survey-weighted univariable and multivariable logistic regression analyses of factors associated with viral load suppression.

Urban residence was associated with lower odds of VLS than rural residence in both the univariable analysis (OR 0.4, 95% CI 0.2–0.7; p=0.0020) and the multivariable analysis (aOR 0.2, 95% CI 0.1–0.5; p=0.0020). Participants who were unaware of their HIV-positive status also had lower odds of VLS than those who were aware of their status (aOR 0.1, 95% CI 0.02–0.1; p<0.001).

There was no evidence of an association between VLS and sex, education level, household wealth quintile, alcohol use, or participation in Teen Club after adjustment.

## Discussion

In this secondary analysis of a nationally representative population-based HIV survey, depressive symptoms were uncommon among adolescents and youth living with HIV (AYLWH) in Malawi and were not significantly associated with viral load suppression (VLS). Six of 201 participants had depressive symptoms, corresponding to a prevalence of 3%. Among the 200 participants with available viral-load results, 137 (68.5%) achieved VLS using the study threshold of <200 copies/mL. In adjusted analyses, urban residence and unawareness of HIV-positive status were associated with lower odds of VLS, whereas sex, education level, household wealth, alcohol use, participation in Teen Club, and depressive symptoms were not significantly associated with VLS.

### Prevalence of depressive symptoms

The prevalence of depressive symptoms observed in this study was substantially lower than estimates reported among adolescents and young people living with HIV elsewhere in sub-Saharan Africa. A systematic review and meta-analysis estimated a prevalence of depression of approximately 26% among adolescents living with HIV in sub-Saharan Africa ^29^. Studies from Kenya and Botswana have reported prevalence estimates of approximately 29% and 23%, respectively ^26,30^. In southern and central Malawi, a study among adolescents living with HIV aged 12–18 years reported a prevalence of depression of approximately 19% ^15^. Differences in study populations, age groups, settings, and instruments used to assess depressive symptoms or depression are likely to contribute to this variation.

The instrument used in the present study is particularly important when interpreting the low prevalence observed. The PHQ-2 is a brief screening instrument rather than a diagnostic assessment, and only six participants screened positive for depressive symptoms. Although the PHQ-2 has demonstrated acceptable performance among adults living with HIV in western Kenya ^27^, it has not been formally validated among Malawian adolescents and youth. The lower prevalence observed in this study could therefore represent a genuinely low prevalence in the surveyed population, limited sensitivity or cultural applicability of the PHQ-2, underreporting, or a combination of these factors. Studies in Malawi have also described limited mental health literacy and stigma surrounding mental illness, which could potentially influence recognition and disclosure of depressive symptoms ^20,21^.

### Viral load suppression

Approximately two-thirds of participants with available viral-load results achieved VLS. This remains substantially below optimal levels for HIV treatment programmes and is consistent with evidence that adolescents and young people can experience poorer virological outcomes than older populations ^4,31^. A study among youth receiving HIV care in Lilongwe, Malawi, using the same <200 copies/mL threshold, reported a higher VLS prevalence of approximately 78% ^5^. Differences between that estimate and the prevalence observed in the present study could reflect differences in study population and setting. The Lilongwe study was conducted among young people engaged in care at a specialised HIV treatment facility, whereas MPHIA was a population-based household survey that included young people regardless of their level of engagement with HIV services. More recent evidence from the African Cohort Study, which included adolescents and young adults aged 15–29 years across Kenya, Nigeria, Tanzania, and Uganda reported viral suppression of 83.8%, with clinic site and ART regimen associated with virological outcomes^32^.

These differences should also be interpreted in the context of the study period. MPHIA 2020–2021 overlapped substantially with the COVID-19 pandemic, including a prolonged interruption in survey data collection. Research from Malawi has documented changes in healthcare utilisation during the pandemic ^33^. Disruptions to routine HIV services and treatment access during this period might therefore have contributed to the VLS levels observed, although the present study was not designed to determine the effect of the pandemic on viral suppression.

### Factors associated with viral load suppression

We found no statistically significant association between depressive symptoms and VLS in either univariable or multivariable analysis. This is consistent with findings from southern Malawi and rural South Africa, where studies also did not identify statistically significant associations between depressive symptoms and virological outcomes ^4,6^. In contrast, studies in other settings have reported associations between depression and viral non-suppression. Among adults living with HIV in Tanzania, persistent depression was associated with an increased risk of viral non-suppression among women, while higher depressive symptom scores were associated with viral non-suppression among men ^11^. A cohort study among adolescents and adults living with HIV in South Africa also reported a higher risk of viral non-suppression among participants with depression ^10^.

Differences in study design, population characteristics, depression measures, viral-load thresholds, and healthcare settings might partly explain these contrasting findings. Importantly, the absence of a statistically significant association in our analysis should not be interpreted as evidence that depressive symptoms have no relationship with VLS. Only six participants screened positive for depressive symptoms, resulting in limited statistical power and imprecise estimates. The adjusted OR of 0.3 had a wide 95% CI of 0.1–2.4, indicating substantial uncertainty around the estimated association. Larger studies using culturally appropriate and more comprehensive mental health assessments are needed to examine this relationship more precisely among AYLWH in Malawi.

Urban residence was independently associated with substantially lower odds of VLS than rural residence. This finding is consistent with a Malawian study of children and adults receiving ART in 2021, which also reported differences in viral suppression between rural and urban populations ^25^. Although urban areas generally have a greater concentration of healthcare facilities, access to and continuity of healthcare can vary considerably within urban settings. A study examining rural–urban differences in access to health services in Malawi demonstrated important heterogeneity within urban areas, including differences between major metropolitan areas and urban townships in the availability of comprehensive primary healthcare services ^23^.

These contextual factors could potentially contribute to the association observed in our study. For example, economic insecurity, mobility, service congestion, stigma, and difficulties accessing youth-friendly services could influence treatment engagement among young people living in urban settings. However, these mechanisms were not directly measured in MPHIA and should therefore be considered possible explanations rather than established causes of the observed association. Further research specifically examining barriers to ART adherence and continuity of HIV care among urban AYLWH in Malawi could help explain this finding.

Awareness of HIV-positive status showed the strongest association with VLS. Approximately three-quarters of participants were aware of their HIV-positive status before receiving the survey test result, whereas nearly one-quarter were unaware. Participants who were unaware of their HIV-positive status had substantially lower odds of VLS than those who were aware. This finding is consistent with a study among adolescents with common mental disorder symptoms in Zimbabwe, which also reported greater odds of virological non-suppression among participants who were unaware of their HIV-positive status ^34^.

The association is also clinically plausible because knowledge of HIV-positive status is a prerequisite for linkage to HIV care and initiation of ART. However, awareness of status should not be interpreted as a direct causal determinant of VLS on the basis of this cross-sectional analysis. Rather, it probably represents an important marker within the pathway from HIV diagnosis to linkage, ART initiation, treatment engagement, and ultimately viral suppression. The finding reinforces the importance of HIV testing, timely communication of results, linkage to care, and sustained treatment engagement among adolescents and youth.

### Strengths and limitations

An important strength of this study is the use of data from MPHIA 2020–2021, a nationally representative population-based HIV survey. The analysis accounted for the complex survey design, including sampling weights, strata, and clusters. Viral load was objectively measured rather than self-reported, and the survey collected a range of demographic, behavioural, psychosocial, and HIV-related variables that could be examined in relation to VLS.

Several limitations should nevertheless be considered. First, the analytical sample was small, comprising 201 AYLWH overall and 200 participants with available viral-load results. Most importantly, only six participants screened positive for depressive symptoms. The resulting low number of exposed participants substantially limited statistical power and precision and might have reduced the ability to detect an association between depressive symptoms and VLS.

Second, depressive symptoms were assessed using the PHQ-2. Although useful as a brief screening instrument, the PHQ-2 is not a diagnostic tool and has not been formally validated among Malawian adolescents and youth. Its brevity could have resulted in misclassification or under-identification of depressive symptoms. Depressive symptoms can also vary over time, whereas the survey captured symptoms at a single timepoint.

Third, the cross-sectional design prevents determination of temporality and does not permit causal inference. It cannot be established, for example, whether depressive symptoms preceded virological outcomes or whether HIV-related experiences and treatment outcomes contributed to depressive symptoms. Similarly, the associations observed for residence and HIV-status awareness should not be interpreted as causal effects.

Fourth, some variables, including alcohol use and depressive symptoms, were based on self-report and could have been affected by recall or social-desirability bias. The relatively small numbers within some categories also resulted in imprecise estimates and necessitated combining categories for regression analysis. Finally, data collection occurred during the COVID-19 pandemic, when healthcare utilisation and HIV service delivery in Malawi were disrupted^33^, which might have influenced treatment engagement and virological outcomes.

Because MPHIA 2020–2021 was a nationally representative population-based survey, the findings are broadly applicable to adolescents and youth living with HIV in Malawi; however, the small analytical sample and low prevalence of depressive symptoms limit the precision of estimates for specific subgroups.

## Conclusion

Among adolescents and youth living with HIV in Malawi, 137 of 200 participants with available viral-load results (68.5%) achieved viral load suppression. Depressive symptoms were uncommon and were not significantly associated with viral load suppression, although the small number of participants screening positive and limitations of the PHQ-2 reduced the precision with which this association could be assessed. Urban residence and unawareness of HIV-positive status were associated with lower odds of viral load suppression. These findings highlight the importance of strengthening HIV testing, linkage to care, and sustained treatment engagement among adolescents and youth, while further research using larger samples and culturally appropriate mental health assessment tools is needed to clarify the relationship between depressive symptoms and HIV treatment outcomes in Malawi.

## Data Availability

The de-identified participant-level data used in this study were obtained from the Malawi Population-based HIV Impact Assessment (MPHIA) 2020-2021 and are available to researchers through the PHIA Data Manager, subject to registration and applicable data-access procedures. The authors do not own the data and cannot directly redistribute the participant-level dataset. The secondary-analysis protocol and R code are available from the corresponding author on reasonable request.

## Cntributors

GN conceived the secondary analysis, developed the study methodology, conducted the data analysis, interpreted the findings, and drafted the manuscript. AdV supervised the study, contributed to the methodology and interpretation of the findings, and critically reviewed and revised the manuscript. GN and AdV directly accessed and verified the underlying participant-level data. Both authors had full access to the data, approved the final version of the manuscript, and accept responsibility for the decision to submit the manuscript for publication.

## Data sharing

The de-identified participant-level data used in this study were obtained from the Malawi Population-based HIV Impact Assessment (MPHIA) 2020–2021 and are not owned by the authors. The MPHIA 2020–2021 household interview and biomarker datasets and intermediary weights are available to researchers through the PHIA Data Manager, subject to registration and the applicable PHIA data-access procedures. The authors therefore cannot directly redistribute the participant-level dataset. The secondary-analysis protocol and R code used for this study are available from the corresponding author on reasonable request following publication.

## Acknowledgments

We thank Annibale Cois for his advice and assistance with the statistical analysis. We also acknowledge ICAP at Columbia University for making the MPHIA 2020–2021 datasets available through the PHIA Data Manager.

## Declaration of generative AI and AI-assisted technologies in the writing process

During preparation of this manuscript, the authors used ChatGPT (OpenAI; GPT-5.6 Sol) to assist with proofreading, punctuation, improving clarity and organisation of manuscript sections. The authors critically reviewed, verified, and edited all AI-assisted output and take full responsibility for the content of the manuscript. The AI tool was not used to generate or analyse participant-level data or to perform the statistical analyses reported in this study.

